# Clinical features, diagnostics, and outcomes of patients presenting with acute respiratory illness: a comparison of patients with and without COVID-19

**DOI:** 10.1101/2020.05.02.20082461

**Authors:** Sachin J. Shah, Peter N. Barish, Priya A. Prasad, Amy Kistler, Norma Neff, Jack Kamm, Lucy M. Li, Charles Y. Chiu, Jennifer M. Babik, Margaret C. Fang, Kirsten Neudoerffer Kangelaris, Charles Langelier, the UCSF COVID-19 Hospital Translational and Clinical Epidemiology Working Group, Yumiko Abe-Jones, Narges Alipanah, Francisco N. Alvarez, Olga Borisovna Botvinnik, Gloria Castaneda, The CZB CLIAhub Consortium, Rand M. Dadasovich, Jennifer Davis, Xianding Deng, Joseph L. DeRisi, Angela M. Detweiler, Scot Federman, John Haliburton, Samantha Hao, Andrew D. Kerkhoff, G. Renuka Kumar, Katherine B. Malcolm, Sabrina A. Mann, Sandra Martinez, Rupa K. Marya, Eran Mick, Lusajo Mwakibete, Nader Najafi, Michael J. Peluso, Maira Phelps, Angela Oliveira Pisco, Kalani Ratnasiri, Luis A. Rubio, Anna Sellas, Kyla D. Sherwood, Jonathan Sheu, Natasha Spottiswoode, Michelle Tan, Guixia Yu

## Abstract

**Background:** Emerging data on the clinical presentation, diagnostics, and outcomes of patients with COVID-19 have largely been presented as case series. Few studies have compared these clinical features and outcomes of COVID-19 to other acute respiratory illnesses.

**Methods:** We examined all patients presenting to an emergency department in San Francisco, California between February 3 and March 31, 2020 with an acute respiratory illness who were tested for SARS-CoV-2. We determined COVID-19 status by PCR and metagenomic next generation sequencing (mNGS). We compared demographics, comorbidities, symptoms, vital signs, and laboratory results including viral diagnostics using PCR and mNGS. Among those hospitalized, we determined differences in treatment (antibiotics, antivirals, respiratory support) and outcomes (ICU admission, ICU interventions, acute respiratory distress syndrome, cardiac injury).

**Findings:** In a cohort of 316 patients, 33 (10%) tested positive for SARS-CoV-2; 31 patients, all without COVID-19, tested positive for another respiratory virus (16%). Among patients with additional viral testing, no co-infections with SARS-CoV-2 were identified by PCR or mNGS. Patients with COVID-19 reported longer symptoms duration (median 7 vs. 3 days) and were more likely to report fever (82% vs. 44%) fatigue (85% vs. 50%) and myalgias (61% vs 27%); p<0.001 for all comparisons. Lymphopenia (55% vs 34%, p=0.018) and bilateral opacities on initial chest radiograph (55% vs. 24%, p=0.001) were more common in patients with COVID-19. Patients with COVID-19 were more often hospitalized (79% vs. 56%, p=0.014). Of 186 hospitalized patients, patients with COVID-19 had longer hospitalizations (median 10.7d vs. 4.7d, p<0.001) and were more likely to develop ARDS (23% vs. 3%, p<0.001). Most comorbidities, home medications, signs and symptoms, vital signs, laboratory results, treatment, and outcomes did not differ by COVID-19 status.

**Interpretation:** While we found differences in clinical features of COVID-19 compared to other acute respiratory illnesses, there was significant overlap in presentation and comorbidities. Patients with COVID-19 were more likely to be admitted to the hospital, have longer hospitalizations and develop ARDS, and were unlikely to have co-existent viral infections. These findings enhance understanding of the clinical characteristics of COVID-19 in comparison to other acute respiratory illnesses.

## Introduction

The severe acute respiratory coronavirus 2 (SARS-CoV-2) and its associated clinical disease, COVID-19, led to a global pandemic in early 2020, with more than 3 million cases and more than 200,000 deaths as of April 2020.^1^ The initial published reports of COVID-19 describe the most common presenting symptoms as fever, cough, and dyspnea.^2–6^ While many people recovered, reports from China, Italy, and the United States showed that approximately 5% of patients required intensive care, and 1.7 to 7.2% died.^1,7,8^ The majority of clinical and outcomes data on COVID-19 have been from Asia and Europe,^4,6,7,9–14^ although data are now emerging from the United States. In particular, studies have reported the clinical features and outcomes of hospitalized patients in Seattle, New York City, and Northern California.^15–19^ However, reports have predominantly focused on patients diagnosed with COVID-19 and have not described in detail the presentation of patients with acute respiratory illness who did not have COVID-19. Without control patients, it is uncertain whether COVID-19 presents differently from other respiratory infections.

The prevalence of viral co-infections in patients with COVID-19 appears to be low in most but not all studies.^15–18,20–23^ However, these studies used conventional microbiological techniques to evaluate for co-infections that are limited in their ability to diagnose respiratory infections.^24^ Understanding the true scope of co-infections in patients with COVID-19 is critical to pursue appropriate diagnostics and management. Metagenomic next-generation sequencing (mNGS) offers a powerful alternative to test for viruses in a respiratory sample in an unbiased manner.^25^

Here we report the clinical characteristics, diagnostics, and outcomes of all patients presenting with respiratory illness to a tertiary academic medical center in San Francisco at the outset of the COVID-19 pandemic. We compare patients with COVID-19 disease to patients presenting during the same time period with an acute respiratory illness and report the prevalence of viral respiratory infections using both conventional microbiology and mNGS.

## Methods

### Setting and design

We conducted a retrospective cohort study to describe the characteristics, diagnostics, and outcomes of patients with respiratory illness presenting to the University of California, San Francisco (UCSF) Health Emergency Department (ED) during the COVID-19 outbreak, comparing patients with and without COVID-19 disease. We identified all patients 18 years or older who underwent testing for COVID-19 within 24 hours of presentation to the ED between February 3 and March 31, 2020.

Two physicians blinded to patients’ COVID-19 status, independently reviewed the documented clinical presentation of all patients and included only those who presented with acute respiratory symptoms (e.g., cough, dyspnea) or influenza-like illness symptoms (e.g., fever, myalgias). Discordant results were re-reviewed together and a consensus decision was reached on all cases (Appendix Figure 1). If patients had multiple encounters during the time period, the first encounter was examined. Patients who were discharged and readmitted within 48 hours were considered a single clinical encounter and outcomes ascertained throughout the encounter.

### Patient characteristics

Patient medical records were reviewed by trained physician chart reviewers and relevant data on initial presentation, radiology findings, and outcomes were abstracted using standardized case review forms. Additional information on patient demographics, vital signs, and laboratory results were obtained from the Epic-based electronic health record. We characterized patients’ comorbidities and their presenting signs and symptoms based on the admission History & Physical and Emergency Department documentation. If a specific comorbidity was not mentioned in the admission documentation, it was considered not present. Records were also reviewed to obtain results of laboratory tests and chest imaging reports within the first 24 hours after admission.

### Clinical microbiological testing

Clinician-ordered testing for COVID-19 was carried out at the UCSF Clinical Microbiology Laboratory by performing reverse transcriptase polymerase chain reaction (PCR) on RNA extracted from oropharyngeal and/or nasopharyngeal swab specimens using primers targeting the SARS-CoV-2 N gene. At the time of the study, PCR results were available at the earliest within 3 hours, and the median time to result was 16 hours. Twenty-six (8%) of the patients had SARS-CoV-2 PCR testing performed at other institutions using their clinically validated assays. Conventional testing for other respiratory viruses was carried out on 270/316 (85%) of patients. This was performed using a 12-target respiratory viral PCR assay (adenovirus, influenza AH1/AH3/B, human metapneumovirus, human rhinovirus, parainfluenza viruses 1–4, respiratory syncytial viruses A/B) or a 3-target (influenza A/B, respiratory syncytial virus PCR) at the discretion of treating clinicians. Bacterial and fungal respiratory pathogens were assessed by semi-quantitative cultures. Patient blood cultures were performed via inoculation into BD Bactec Plus Aerobic and Lytic Anaerobic media (Becton Dickinson).

### Respiratory virus detection by metagenomic sequencing

To further screen for the presence of other respiratory viral pathogens, metagenomic next generation sequencing (mNGS) of RNA was performed on available residual RNA extracted for COVID-19 clinical PCR testing on 107 randomly selected patients. After DNase treatment, human ribosomal RNA depletion was carried out using FastSelect (Qiagen). To control for background contamination, we included negative controls (water and HeLa cell RNA) as well as positive controls (spike-in dilution series of RNA standards from the External RNA Controls Consortium [ERCC]).^26^ The latter enabled subsequent bioinformatic assessment of the total RNA mass input in each sample.^27^

RNA was then fragmented and subjected to a modified metagenomic spiked sequencing primer enrichment (MSSPE) library preparation method.^28^ Briefly, a 1:1 mixture of the NEBNext Ultra II RNAseq Library Prep (New England Biolabs) random primer stock and a pool of SARS-CoV-2 primers at 100 μM was used at the first strand synthesis step of the standard RNAseq library preparation protocol to enrich for the recovery of reads spanning the length of the SARS-CoV-2 genome sequence in the context of mNGS analysis.^29^ RNA-seq libraries underwent 146 nucleotide paired-end Illumina sequencing on an Illumina NovaSeq 6000.

### mNGS bioinformatic and phylogenetic analysis

Following demultiplexing, reads were host- and quality-filtered and then subjected to viral reference based alignment at both the nucleotide and amino acid level against sequences in the National Center for Biotechnology Information (NCBI) nucleotide (NT) and non-redundant (NR) databases, followed by assembly using previously validated bioinformatics pipelines.^30,31^ Samples (n=10) with insufficient input RNA for accurate viral assessment (< 25 pg, calculated based on alignments to positive control ERCC RNA standards) were considered invalid, leaving 97 subjects available for analysis.

Negative control (water and HeLa cell RNA) samples enabled estimating the number of background reads to each virus, which were normalized by input mass determined based on the ratio of sample reads to spike-in positive control ERCC RNA standards.^27^ Viruses with sequencing reads significantly greater compared to negative controls (adjusted p value < 0.05 using a Holm-Bonferroni correction within each sample) were identified by modeling the number of background reads as a negative binomial distribution with mean and dispersion fitted on the negative controls. For phylogenetic analysis of SARS-CoV-2 viruses, we constructed genomes using minimap2^32^ to align reads to the reference MN908947.3 and iVar^33^ to trim primers and call variants, then restricted to samples with at least 10-fold coverage of at least 97% (29 kilobases) of the genome (n=10) and utilized the Nextstrain^34^ pipeline to build a phylogenetic tree using iqtree.^35^ Viral genomic data is publicly accessible via gisaid.org (Global Initiative on Sharing All Influenza Data) ^36^ and Genbank (MT385414 - MT385497).

### Treatment and Outcomes

Clinical treatment and outcomes were ascertained through a combination of chart review and extraction of structured fields from the electronic health record. Medication records were reviewed to identify the administration of relevant antibiotics. We determined if patients required respiratory support at any point during their hospitalization: nasal cannula, high flow nasal cannula, noninvasive ventilation (bilevel or continuous positive airway pressure) or endotracheal intubation. Patients were considered to have new-onset cardiomyopathy if a treating physician documented the diagnosis. Acute respiratory distress syndrome (ARDS) was defined according to the Berlin definition by two physicians.^37^ Acute kidney injury was defined using the Kidney Disease: Improving Global Outcomes definition.^38^ Outcome ascertainment was censored on April 25, 2020.

### Statistical analysis

We used descriptive statistics to characterize the features of patients grouped by COVID infection. Where clinically relevant we dichotomized continuous variables. For normally distributed continuous variables we calculated the mean and standard deviation and tested for differences using t-tests. For non-normally distributed continuous variables we calculated the median and interquartile range and tested for differences using the Wilcoxon rank sum test. For categorical and dichotomous variables we evaluated differences between groups using the chi-square test or Fisher’s exact test. The analyses were not adjusted for multiple comparisons and should be interpreted as descriptive and exploratory. The Human Research Protection Program Institutional Review Board at the University of California, San Francisco, approved this study (IRB# 16-20956). We used Stata version 14.2 (College Station, TX) and SAS version 9.4 (Cary, NC) to conduct all analyses.

## Results

### Demographic characteristics and comorbidities

Out of 316 patients who presented with acute respiratory illness and underwent testing for COVID-19, 33 (10%) tested positive for SARS-CoV-2 by PCR. Patients with a positive COVID-19 test result were more likely to have traveled to an area of community transmission or to have had contact with someone with COVID-19 (46% vs 11%, p<0.001) to be married (64% vs. 36%, p = 0.02) or to identify as Asian (42% vs. 24%, p= 0.010) (**Table 1**). Patients who tested positive were also more likely to report never smoking tobacco (61% vs. 40%, p=0.001) and to have undergone solid organ transplantation (12% vs. 3%, p=0.027). The prevalence of hypertension and diabetes did not differ significantly between COVID-19 positive and negative patients. There was no significant difference by COVID-19 status of the proportion of patients taking an angiotensin-converting enzyme inhibitor or angiotensin II receptor blocker.

**Table 1:**
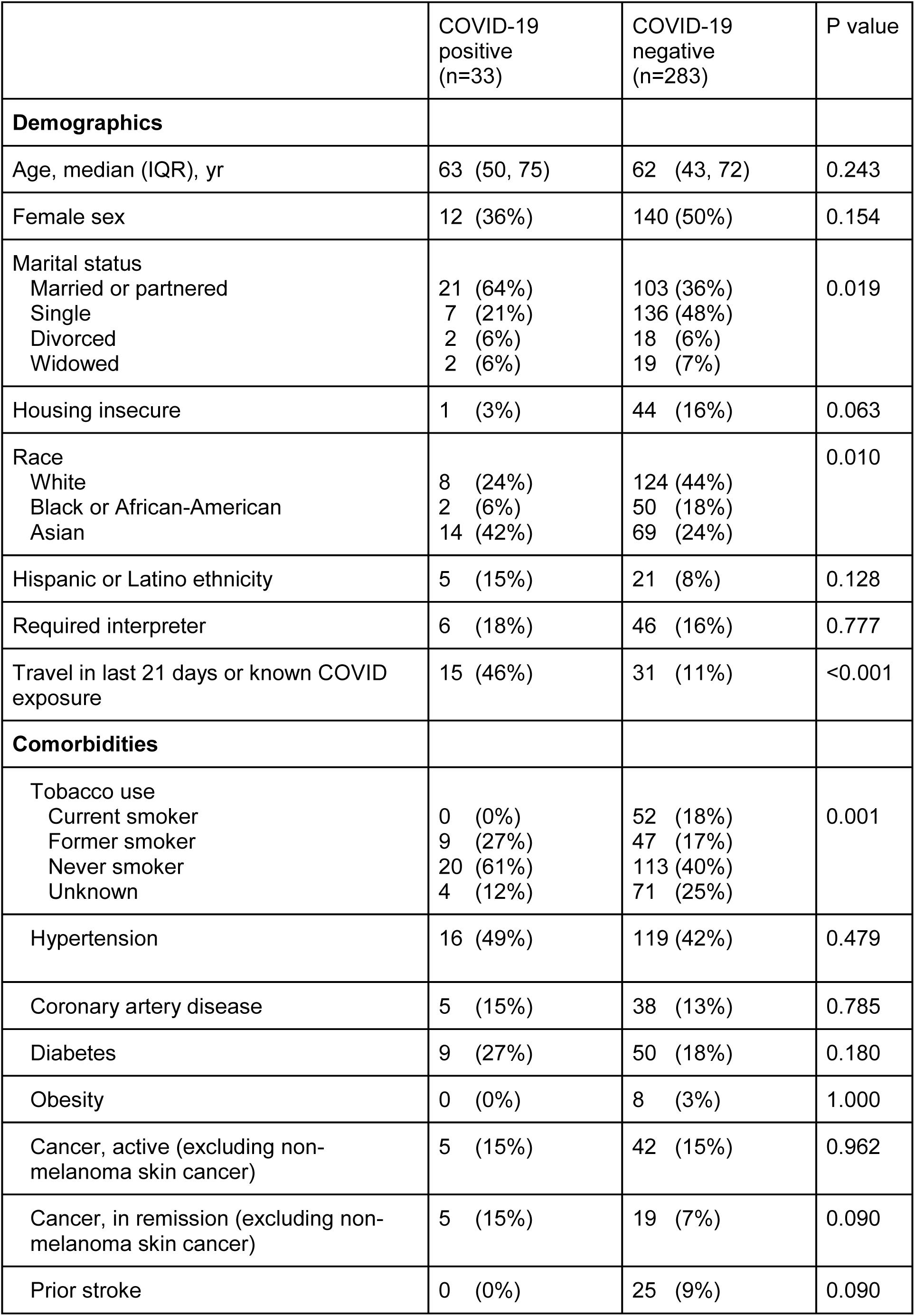
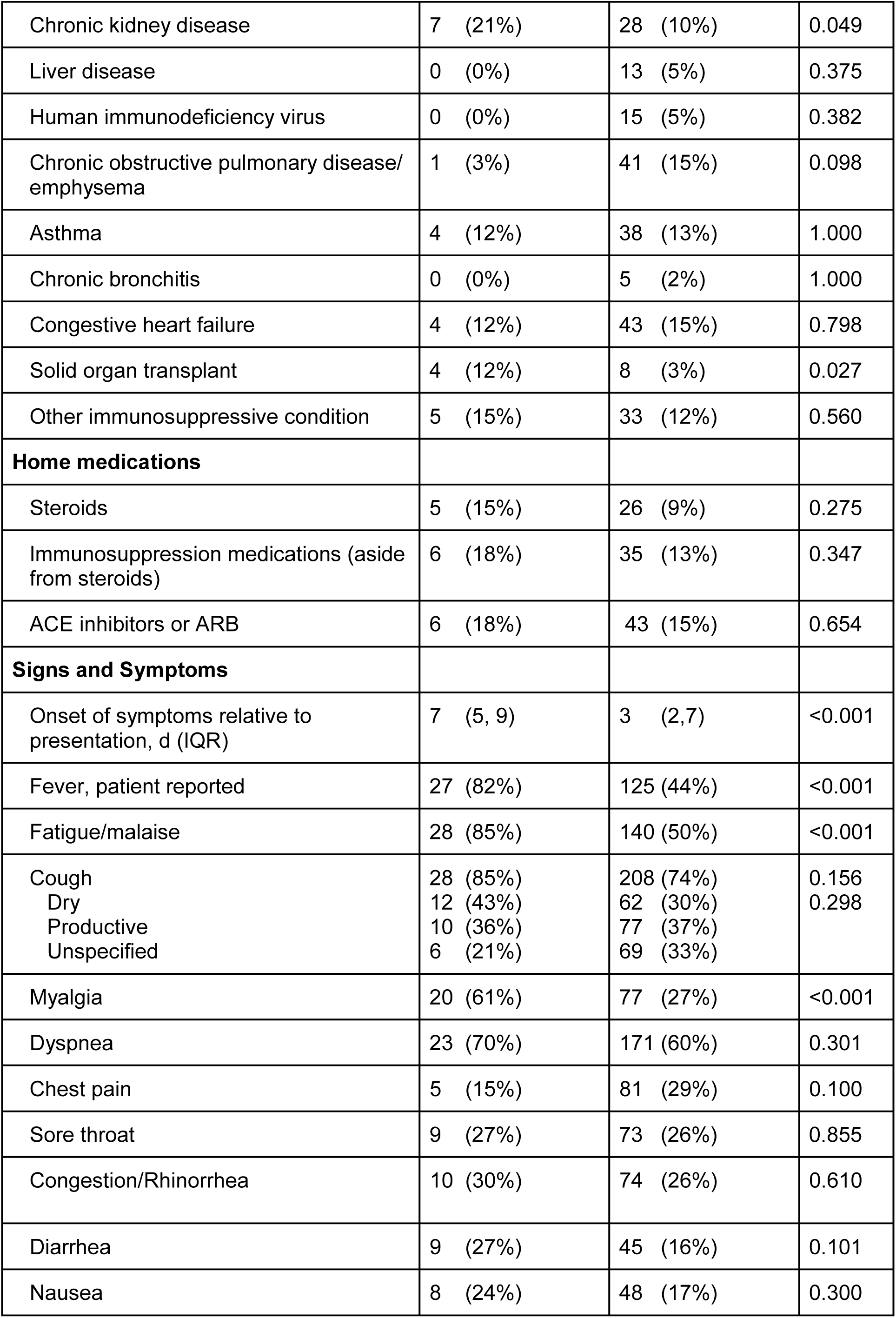
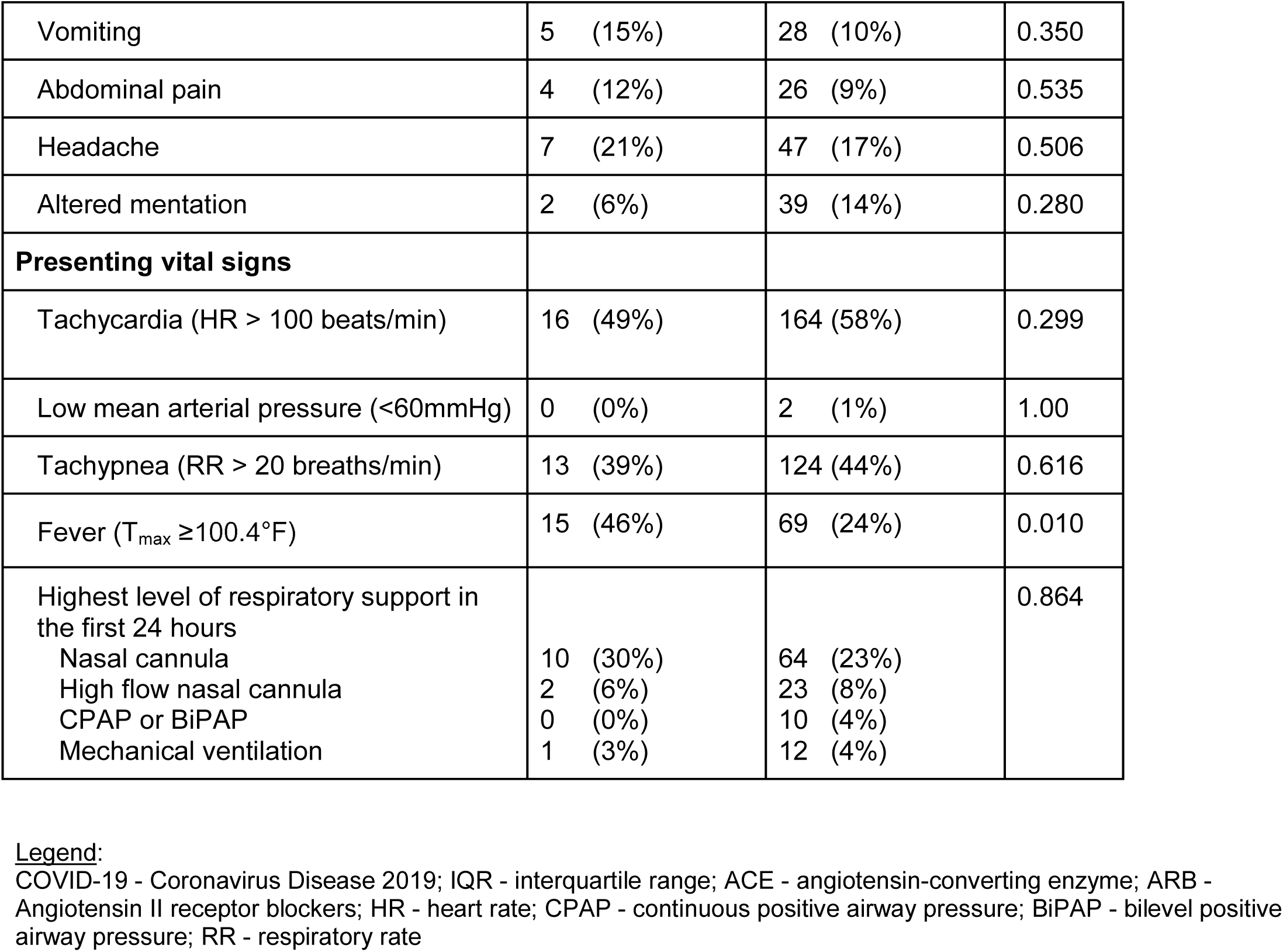
Characteristics of 316 patients presenting with acute respiratory illness and tested for COVID-19

**Table 2:**
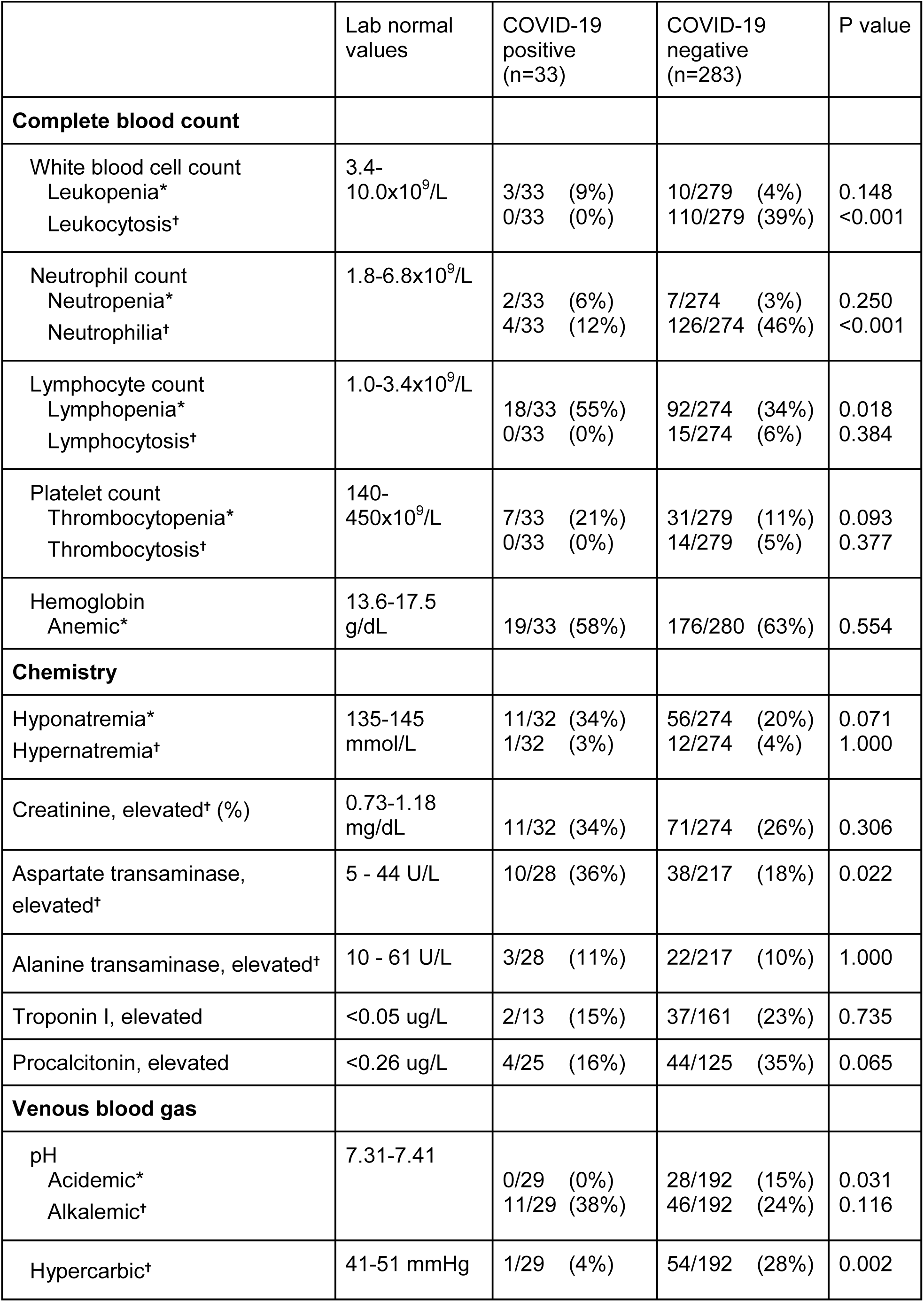
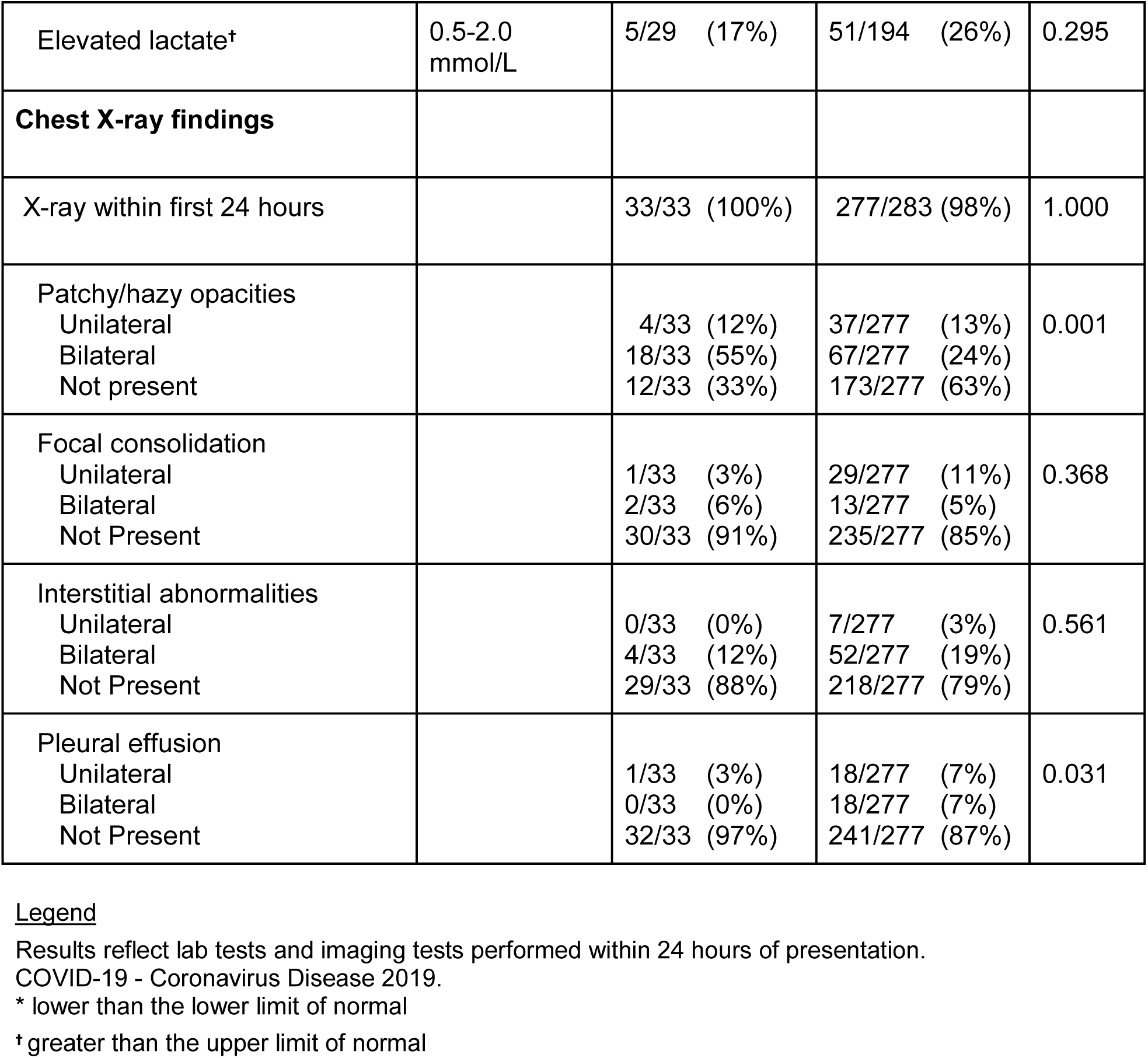
Laboratory and imaging findings within 24 hours of presentation among 316 patients presenting with acute respiratory illness and tested for COVID-19

### Signs, symptoms and vital signs

Patients with COVID-19 reported a longer duration of symptoms prior to ED presentation (median 7 vs. 3 days, p<0.001) (**Table 1**). COVID-19 patients reported fever (82% vs. 44%, p<0.001) fatigue (85% vs. 50%, p<0.001) and myalgias (61% vs 27%, p<0.001) at a higher rate than COVID-19 negative patients. The presence and characteristics of cough, dyspnea, and chest pain did not differ based on COVID-19 infection. Gastrointestinal symptoms -- nausea, vomiting, diarrhea, and abdominal pain -- were present at similar rates in the two groups. With respect to vital sign abnormalities, tachycardia, hypotension, oxygen requirement, and tachypnea did not differ by COVID-19 status. However, patients with COVID-19 were more likely to present with a measured fever (46% vs 24%, p=0.010).

### Laboratory studies and imaging upon presentation

Lymphopenia was more common in patients with COVID-19 at the time of presentation (55% vs 34%, p=0.018). Aspartate transaminase but not alanine transaminase was more often elevated in patients with COVID-19 (36% vs. 18% p=0.022 and 11% vs. 10% p=1.000, respectively). Patients with COVID-19 were less often acidemic (0% vs. 15%, p=0.031) and less often found to be hypercarbic (4% vs. 28%, p=0.002) by venous blood gas. Of the patients tested on presentation, neither troponin nor procalcitonin elevation differed by COVID-19 status. Chest X-rays were performed on all but 6 patients. Radiographs from patients with COVID-19 were more likely to reveal bilateral patchy or hazy opacities (55% vs. 24%, p=0.001). Focal consolidations, interstitial abnormalities, and pleural effusions were observed at similar proportions.

### Pathogen diagnostics

Clinicians ordered Influenza/Respiratory syncytial virus PCR testing for 99/316 (31%) patients and 12-target respiratory virus PCR for 171/316 (54%) patients; testing rates did not differ by COVID-19 status (**Table 3**). Orthogonal mNGS analysis was performed on swab specimens from 97/316 (31%) of patients to provide additional broad range screening of both common and uncommon viral pathogens. By PCR, SARS-CoV-2 was the most prevalent respiratory virus detected, in 33/316 patients (10%). No co-infections with SARS-CoV-2 and other viruses were identified. Other respiratory viruses were identified in 31/194 (16%) of patients without COVID-19. Independent mNGS analyses corroborated 13/14 (93%) of SARS-CoV-2 infections and 11/11 (100%) of other respiratory viral infections detected by clinical PCR assays. Respiratory bacterial co-infection was not more common in patients with COVID-19 (11% vs. 18%, p=1.000) and no cases of ventilator associated pneumonia were identified in COVID-19 patients. Bacteremia or fungemia was also not more common in patients with COVID-19 disease (5% vs. 7%, p =1.00).

**Table 3:**
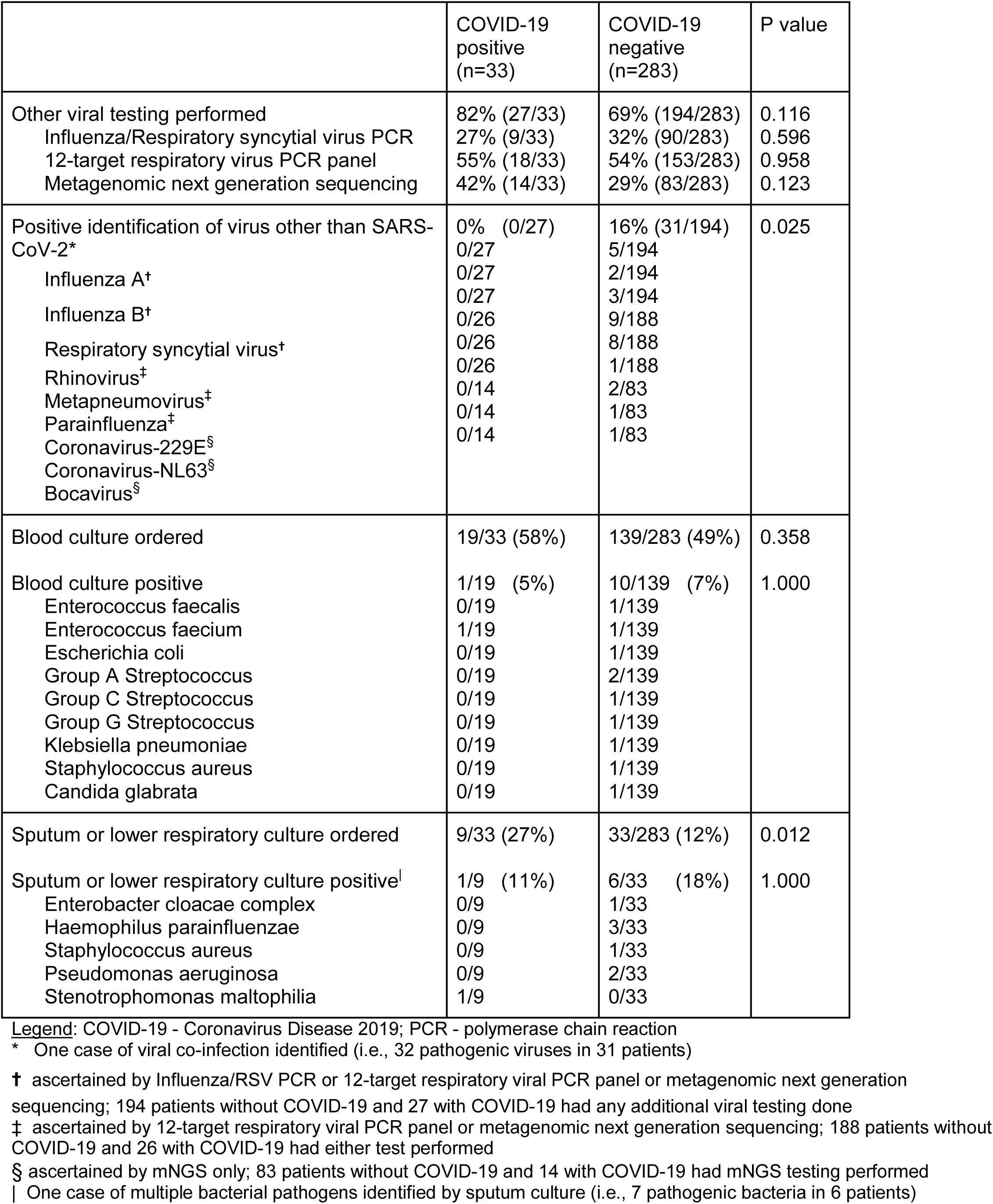
Results of infectious disease testing among 316 patients presenting with acute respiratory illness and tested for COVID-19

|  | COVID-19 positive (n=33) | COVID-19 negative (n=283) | P value |
| --- | --- | --- | --- |
| Other viral testing performed | 82% (27/33) | 69% (194/283) | 0.116 |
| Influenza/Respiratory syncytial virus PCR | 27% (9/33) | 32% (90/283) | 0.596 |
| 12-target respiratory virus PCR panel | 55% (18/33) | 54% (153/283) | 0.958 |
| Metagenomic next generation sequencing | 42% (14/33) | 29% (83/283) | 0.123 |
| Positive identification of virus other than SARS-CoV-2* | 0% (0/27) | 16% (31/194) | 0.025 |
| Influenza A<sup>†</sup> | 0/27 | 5/194 |  |
| Influenza B<sup>†</sup> | 0/27 | 2/194 |  |
| Respiratory syncytial virus<sup>†</sup> | 0/27 | 3/194 |  |
| Rhinovirus<sup>‡</sup> | 0/26 | 9/188 |  |
| Metapneumovirus<sup>‡</sup> | 0/26 | 8/188 |  |
| Parainfluenza<sup>‡</sup> | 0/26 | 1/188 |  |
| Coronavirus-229E<sup>§</sup> | 0/14 | 2/83 |  |
| Coronavirus-NL63<sup>§</sup> | 0/14 | 1/83 |  |
| Bocavirus<sup>§</sup> | 0/14 | 1/83 |  |
| Blood culture ordered | 19/33 (58%) | 139/283 (49%) | 0.358 |
| Blood culture positive | 1/19 (5%) | 10/139 (7%) | 1.000 |
| Enterococcus faecalis | 0/19 | 1/139 |  |
| Enterococcus faecium | 1/19 | 1/139 |  |
| Escherichia coli | 0/19 | 1/139 |  |
| Group A Streptococcus | 0/19 | 2/139 |  |
| Group C Streptococcus | 0/19 | 1/139 |  |
| Group G Streptococcus | 0/19 | 1/139 |  |
| Klebsiella pneumoniae | 0/19 | 1/139 |  |
| Staphylococcus aureus | 0/19 | 1/139 |  |
| Candida glabrata | 0/19 | 1/139 |  |
| Sputum or lower respiratory culture ordered | 9/33 (27%) | 33/283 (12%) | 0.012 |
| Sputum or lower respiratory culture positive | 1/9 (11%) | 6/33 (18%) | 1.000 |
| Enterobacter cloacae complex | 0/9 | 1/33 |  |
| Haemophilus parainfluenzae | 0/9 | 3/33 |  |
| Staphylococcus aureus | 0/9 | 1/33 |  |
| Pseudomonas aeruginosa | 0/9 | 2/33 |  |
| Stenotrophomonas maltophilia | 1/9 | 0/33 |  |

Legend: COVID-19 - Coronavirus Disease 2019; PCR - polymerase chain reaction
\* One case of viral co-infection identified (i.e., 32 pathogenic viruses in 31 patients)
† ascertained by Influenza/RSV PCR or 12-target respiratory viral PCR panel or metagenomic next generation sequencing; 194 patients without COVID-19 and 27 with COVID-19 had any additional viral testing done
‡ ascertained by 12-target respiratory viral PCR panel or metagenomic next generation sequencing; 188 patients without COVID-19 and 26 with COVID-19 had either test performed
§ ascertained by mNGS only; 83 patients without COVID-19 and 14 with COVID-19 had mNGS testing performed
| One case of multiple bacterial pathogens identified by sputum culture (i.e., 7 pathogenic bacteria in 6 patients)

### Genomic epidemiology of SARS-CoV-2

To understand the genomic epidemiology of SARS-CoV-2 in the cohort, phylogenetic analysis was performed. SARS-CoV-2 genomes with at least 97% coverage at 10-fold sequencing depth could be recovered from 10 of the 13 mNGS-positive subjects. These 10 genomes originate from several parts of the global SARS-CoV-2 phylogeny, with clades A2a (n=3, widely prevalent in New York) and B1 (n=3, detected in Washington State in February 2020) representing slightly more than half of the lineages we identified (**Appendix Figure 2**). The SARS-CoV-2 isolated from patients who required ICU care were not associated with any single clade.

### Hospitalization treatment and outcomes

In all, 186 patients were hospitalized and patients with COVID-19 were more likely to be admitted (79% vs. 56%, p=0.014) and have longer lengths of stay (median 10.7 vs. 4.7 days, p<0.001). Among hospitalized patients, antibiotics and oseltamivir were used in similar proportions (**Table 4**). Hydroxychloroquine was more often used in patients with COVID-19 (22% vs. < 1%, p<0.001); however, azithromycin and corticosteroids use did not differ by COVID-19 status. Six of 26 inpatients with COVID-19 were enrolled in a randomized trial of remdesivir. Respiratory support was provided in similar proportions of patients and, when respiratory support was needed, the level of support did not differ by COVID-19 status.

**Table 4:** Treatment of 186 hospitalized patients with acute respiratory illness and tested for COVID-19

|  | COVID-19 positive<br>(n=26) | COVID-19<br>negative<br>(n=160) | P value |
| --- | --- | --- | --- |
| Antibiotics administered | 17/26 (65%) | 134/160 (84%) | 0.054 |
| Vancomycin | 8/26 (31%) | 72/160 (45%) | 0.126 |
| Piperacillin/tazobactam | 5/26 (19%) | 55/160 (35%) | 0.107 |
| Cefepime | 4/26 (15%) | 17/160 (11%) | 0.504 |
| Ceftriazone | 10/26 (39%) | 74/160 (46%) | 0.459 |
| Carbapenems | 3/26 (12%) | 19/160 (12%) | 1.000 |
| Azithromycin | 8/26 (31%) | 44/160 (28%) | 0.731 |
| Doxycycline | 7/26 (29%) | 70/160 (44%) | 0.106 |
| Fluoroquinolones | 4/26 (15%) | 32/160 (20%) | 0.581 |
| Other antibiotics | 4/26 (15%) | 43/160 (27%) | 0.329 |
| Oseltamivir | 3/26 (12%) | 15/160 (9%) | 0.729 |
| Remdesivir clinical trial* | 6/26 (23%) | 0/160 (0%) | <0.001 |
| Chloroquine | 0/26 (0%) | 0/160 (0%) | --- |
| Hydroxychloroquine | 6/26 (22%) | 1/160 (<1%) | <0.001 |
| Steroids | 3/26 (12%) | 23/160 (14%) | 1.000 |
| No respiratory support | 6/26 (23%) | 55/160 (34%) | 0.255 |
| Respiratory support |  |  |  |
| Supplemental oxygen | 10/20 (50%) | 61/105 (58%) | 0.711 |
| High flow oxygen | 5/20 (25%) | 21/105 (20%) |  |
| Noninvasive positive-pressure<br>ventilation or invasive<br>mechanical ventilation | 5/20 (25%) | 23/105 (22%) |  |

COVID-19 - Coronavirus Disease 2019
\* Rows are not mutually exclusive, 1 patient received hydroxychloroquine and was enrolled in a blinded remdesivir trial

Numerically, more patients with COVID-19 required ICU care compared to non-COVID-19 patients, although the difference was not statistically significant (42% vs. 26%, p=0.092) (**Table 5**). When transferred to the ICU, there was no observed difference in the use of ICU interventions; however, patients with COVID-19 had a longer ICU length of stay (median 8.8 vs. 2.9 days, p=0.005). Those diagnosed with COVID-19 were more likely to develop ARDS (23% vs. 3%, p<0.001) but were no more likely to develop cardiomyopathy or acute kidney injury when compared to non-COVID-19 patients. Among those tested, patients diagnosed with COVID-19 were no more often observed to have abnormal coagulation tests or elevated troponin.

**Table 5:**
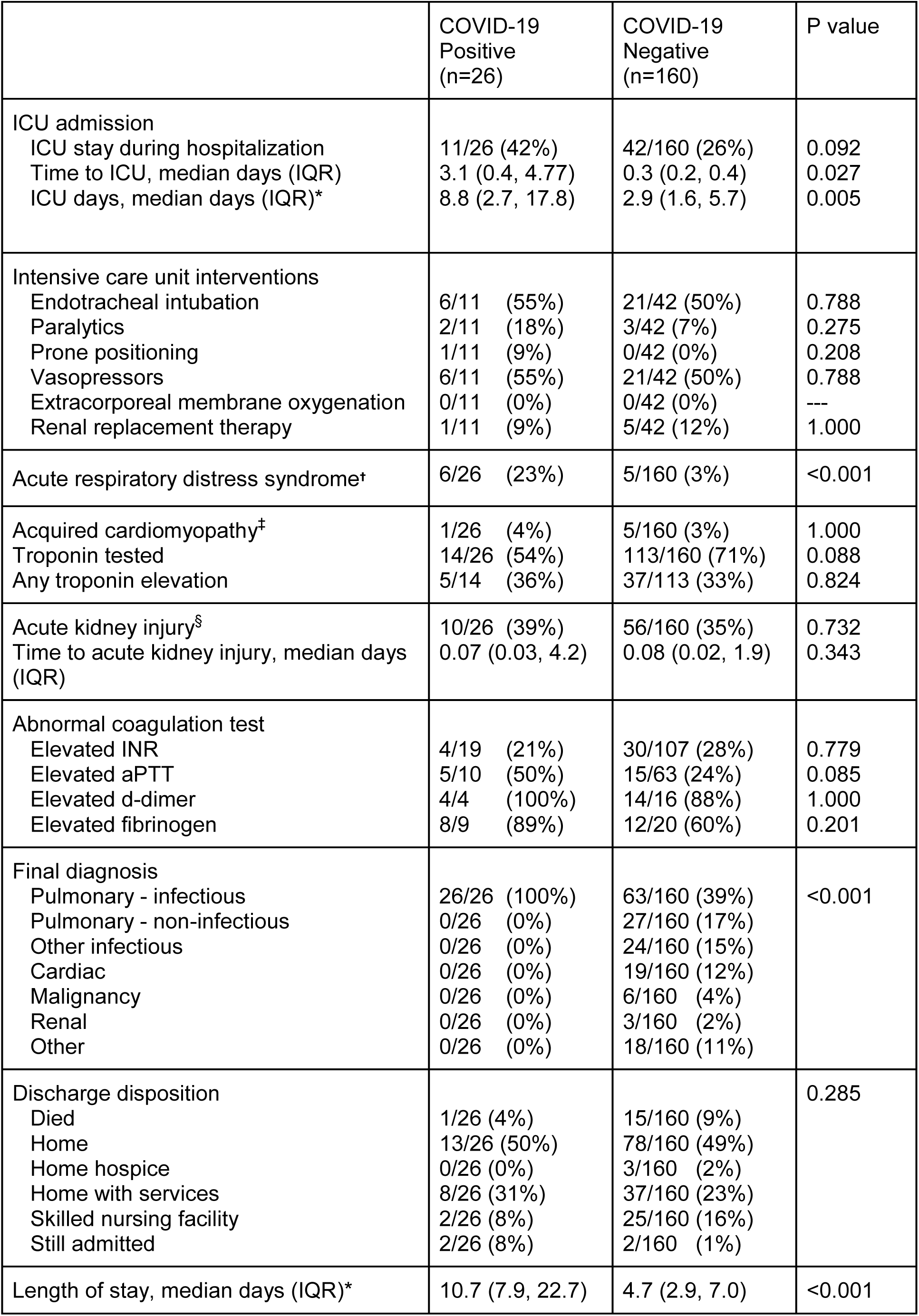
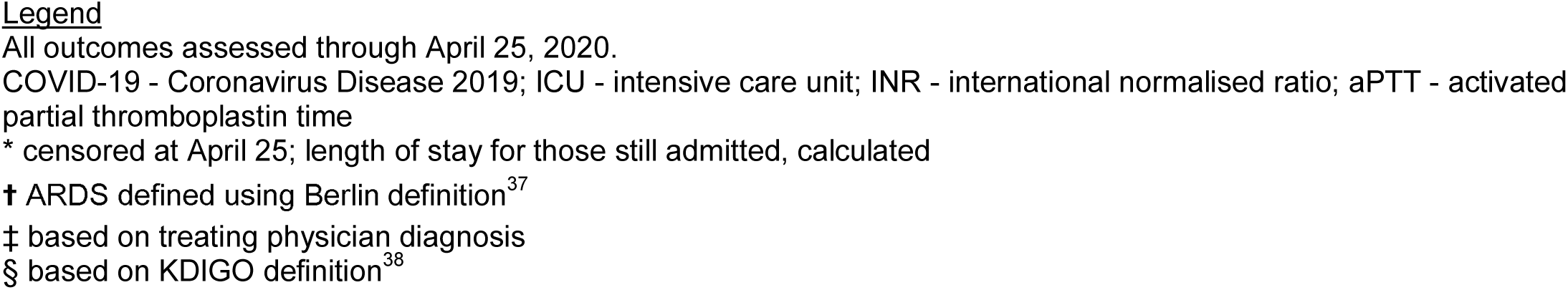
Outcomes of 186 hospitalized patients with acute respiratory illness and tested for COVID-19

|  | COVID-19<br>Positive<br>(n=26) | COVID-19<br>Negative<br>(n=160) | P value |
| --- | --- | --- | --- |
| ICU admission |  |  |  |
| ICU stay during hospitalization | 11/26 (42%) | 42/160 (26%) | 0.092 |
| Time to ICU, median days (IQR) | 3.1 (0.4, 4.77) | 0.3 (0.2, 0.4) | 0.027 |
| ICU days, median days (IQR)* | 8.8 (2.7, 17.8) | 2.9 (1.6, 5.7) | 0.005 |
| Intensive care unit interventions |  |  |  |
| Endotracheal intubation | 6/11 (55%) | 21/42 (50%) | 0.788 |
| Paralytics | 2/11 (18%) | 3/42 (7%) | 0.275 |
| Prone positioning | 1/11 (9%) | 0/42 (0%) | 0.208 |
| Vasopressors | 6/11 (55%) | 21/42 (50%) | 0.788 |
| Extracorporeal membrane oxygenation | 0/11 (0%) | 0/42 (0%) | --- |
| Renal replacement therapy | 1/11 (9%) | 5/42 (12%) | 1.000 |
| Acute respiratory distress syndrome† | 6/26 (23%) | 5/160 (3%) | <0.001 |
| Acquired cardiomyopathy‡ | 1/26 (4%) | 5/160 (3%) | 1.000 |
| Troponin tested | 14/26 (54%) | 113/160 (71%) | 0.088 |
| Any troponin elevation | 5/14 (36%) | 37/113 (33%) | 0.824 |
| Acute kidney injury§ | 10/26 (39%) | 56/160 (35%) | 0.732 |
| Time to acute kidney injury, median days (IQR) | 0.07 (0.03, 4.2) | 0.08 (0.02, 1.9) | 0.343 |
| Abnormal coagulation test |  |  |  |
| Elevated INR | 4/19 (21%) | 30/107 (28%) | 0.779 |
| Elevated aPTT | 5/10 (50%) | 15/63 (24%) | 0.085 |
| Elevated d-dimer | 4/4 (100%) | 14/16 (88%) | 1.000 |
| Elevated fibrinogen | 8/9 (89%) | 12/20 (60%) | 0.201 |
| Final diagnosis |  |  |  |
| Pulmonary - infectious | 26/26 (100%) | 63/160 (39%) | <0.001 |
| Pulmonary - non-infectious | 0/26 (0%) | 27/160 (17%) |  |
| Other infectious | 0/26 (0%) | 24/160 (15%) |  |
| Cardiac | 0/26 (0%) | 19/160 (12%) |  |
| Malignancy | 0/26 (0%) | 6/160 (4%) |  |
| Renal | 0/26 (0%) | 3/160 (2%) |  |
| Other | 0/26 (0%) | 18/160 (11%) |  |
| Discharge disposition |  |  | 0.285 |
| Died | 1/26 (4%) | 15/160 (9%) |  |
| Home | 13/26 (50%) | 78/160 (49%) |  |
| Home hospice | 0/26 (0%) | 3/160 (2%) |  |
| Home with services | 8/26 (31%) | 37/160 (23%) |  |
| Skilled nursing facility | 2/26 (8%) | 25/160 (16%) |  |
| Still admitted | 2/26 (8%) | 2/160 (1%) |  |
| Length of stay, median days (IQR)* | 10.7 (7.9, 22.7) | 4.7 (2.9, 7.0) | <0.001 |

## Discussion

While a number of studies describe the clinical features of patients with COVID-19, few have directly compared the clinical presentation and outcomes of COVID-19 to other respiratory illnesses.^23,39–43^ Without a control group, and in settings of restricted COVID-19 test availability, we cannot ascertain whether COVID-19 presents differently from other forms of respiratory illnesses. In our study comparing acutely ill patients with and without COVID-19 presenting for emergency care, we found that patients with COVID-19 had a longer duration of symptoms, were more likely to be admitted to the hospital, had longer hospitalizations and were more likely to develop ARDS. Using standard laboratory PCR testing, and mNGS, we found a 16% prevalence of other respiratory viruses in the COVID-19 negative patients, and a lack of detectable viral co-infections in the COVID-19 positive patients.

Patients diagnosed with COVID-19 were more likely to be Asian (44%) which may reflect differences in the dynamics of infection early in the COVID-19 pandemic as well as the high proportion of people in San Francisco who self-identify as Asian (36%).^44^ COVID-19 patients were more likely to be never smokers, in line with other studies showing no link between tobacco use and increased COVID-19 risk.^4 45,46^ Largely similar comorbidity profiles were observed between COVID-19 positive and negative patients, aside from a higher proportion of chronic kidney disease and history of solid organ transplantation in COVID-19 patients.

Patients diagnosed with COVID-19 had a longer duration of symptoms prior to presentation and were more likely than control patients to report fever, fatigue and myalgias. It is notable, however, that 44% of COVID-19 negative patients reported fevers and systemic symptoms were common. In contrast to other reports,^4,6,7^ COVID-19 positive patients in this cohort had relatively high rates of upper respiratory symptoms (21% with headache, 27% with sore throat, and 30% with congestion/rhinorrhea) and gastrointestinal symptoms. In terms of laboratory values, patients with COVID-19 were significantly more likely to have lymphopenia and no patient with COVID-19 had leukocytosis.

Determining rates of co-infection in patients with COVID-19 has significance given that SARS-CoV-2 testing may be deferred if an alternative respiratory pathogen is identified, especially in settings with limited test availability. In this cohort, no patients with COVID-19 had evidence of viral co-infection, by either clinical PCR testing or by mNGS analysis. Only one COVID-19 positive patient had evidence of co-infection with a bacterial respiratory pathogen, and no difference in the prevalence of bacterial co-infection was identified based on COVID-19 status. These results are distinct from those reported in a recent study of COVID-positive patients that found a 21% rate of viral co-infections^23^ but consistent with data from several other institutions demonstrating very low rates (≤6%) of viral or bacterial co-infection in hospitalized COVID-19 positive patients, including two recent large studies from New York City.^15–18,20–23^ Further investigation of co-infections in COVID-19 positive patients, and assessment of their potential impact on disease severity and outcomes is needed, especially if SARS-CoV-2 circulation extends to overlap with other highly prevalent seasonal respiratory pathogens.

Although patients with COVID-19 were more likely to be diagnosed with ARDS, there were no differences in their need for ICU care or mechanical ventilation. We also did not find significant differences in terms of acquired cardiomyopathy or troponin elevation during the hospitalization. Despite concerns for cardiac complications in COVID-19 positive patients, our findings highlight the importance of comparisons to control groups of hospitalized patients.^16,47,48^ Large proportions of patients in both groups received broad-spectrum antibiotics, despite all of the COVID-19 positive patients having a confirmed viral etiology. This has important implications for antibiotic stewardship in the COVID-19 era and likely reflects clinical uncertainty about the true rate of bacterial co-infection early in the pandemic. COVID-19 was associated with longer hospital lengths of stay. While the duration of hospitalization may reflect the severity of illness, it could also be a marker of concern for late decompensation in these patients^49^ or difficulties with hospital discharge due to requirements for isolation and infection control.

Prior studies describing the clinical presentation of patients with COVID-19 have for the most part identified non-specific features that characterize respiratory infections in general. To our knowledge this is the first U.S. study to identify characteristics distinguishing patients with COVID-19 from patients who underwent investigation for COVID-19 but were ultimately found to have an alternate diagnosis. Previous publications on this topic are primarily smaller in scope and are all outside of the US.^39,40,42^ The clinical, laboratory, and imaging data we highlight have important implications for front line providers making decisions in real-time regarding the pretest probability of COVID-19, especially in settings with limited access to rapid COVID-19 diagnostics.

In contrast to other areas in the United States, the Bay Area has not yet experienced a large surge in cases of COVID-19. The fact that resources were not strained may have affected the clinical course and outcomes observed. For example, while sample size is not sufficient to evaluate differences in mortality, only one of the 33 with COVID-19 died (3%) which is lower than in other studies of hospitalized U.S. patients.^17,18^ There is speculation that variations in circulating SARS-CoV-2 strains may affect pathogenicity and contribute to geographic differences in case fatality rates.^50,51^ Exploratory phylogenetic analysis presented here demonstrated a diversity of strains among the COVID-19 patients requiring ICU care without a predominant clade; larger studies are needed to assess any potential relationship.

There are several limitations inherent to the study design and data available that should be considered when interpreting the results of this study. As a retrospective study based in a single academic medical center and focusing on patients presenting for emergency care, it may not generalize to other institutions with different patient populations or patients with milder forms of disease. Variation in clinician assessment and documentation may lead to misclassification of some variables. Although all patients in the COVID-19 negative group presented with respiratory complaints and/or influenza-like illness, only 56% of patients were given a final diagnosis of respiratory infection, which may affect the generalizability of our outcomes data. Finally, this study was undertaken at the end of the influenza season and during a period of social distancing, both of which likely impacted the prevalence of circulating viruses and the rate of co-infections.

In summary, while many clinical features of COVID-19 overlap with those of other acute respiratory illnesses, several unique characteristics were identified. Patients with COVID-19 had a longer duration of symptoms, particularly fatigue, fever, and myalgias, were more likely to be admitted to the hospital and for a longer duration, were unlikely to have co-existent viral infections, and were more likely to develop ARDS. Though this health system has not experienced a surge in COVID-19 cases, these key clinical characteristics may, in part, explain the observed differences in propensity of COVID-19 to strain health systems. While we did find meaningful differences that may inform one's clinical suspicion for COVID-19, we did not find significant differences in cardiopulmonary comorbidities, ACE inhibitor/ARB use, or mortality rate. These findings enhance understanding of the clinical characteristics of COVID-19 in comparison to other acute respiratory illnesses.

## Data Availability

Viral genomic data is publicly accessible via gisaid.org (Global Initiative on Sharing All Influenza Data) 36 and Genbank (MT385414 - MT385497).

## Author contributions

Drs. Shah and Langelier had full access to all of the data and take responsibility for the integrity of the data and the accuracy of the data analysis.

*Concept and design:* Shah, Barish, Prasad, Kistler, Babik, Fang, Kangelaris, Langelier

*Acquisition, analysis, or interpretation of data:* Shah, Barish, Prasad, Kistler, Kamm, Li, Chiu, Babik, Fang, Kangelaris, Langelier, Abe-Jones, Alipanah, Alvarez, Botvinnik, Castaneda, The CZB CLIAhub Consortium, Dadasovich, Davis, Deng, Detweiler, Federman, Haliburton, Hao, Kerkhoff, Kumar, Malcolm, Mann, Martinez, Marya, Mick, Mwakibete, Najafi, Peluso, Phelps, Pisco, Ratnasiri, Rubio, Sellas, Sherwood, Spottiswoode, Tan, Yu

*Drafting of the manuscript:* Shah, Barish, Kistler, Kamm, Babik, Fang, Kangelaris, Langelier

*Critical revision of the manuscript for important intellectual content:* All authors

*Statistical analysis:* Shah, Prasad, Li, Kamm, Hao, Martinez

*Obtained funding:* Shah, Chiu, Fang, Kangelaris, Langelier, DeRisi

*Supervision:* Shah, Kistler, Chiu, Kangelaris, Langelier, DeRisi

## Appendix

**Appendix Figure 1:**
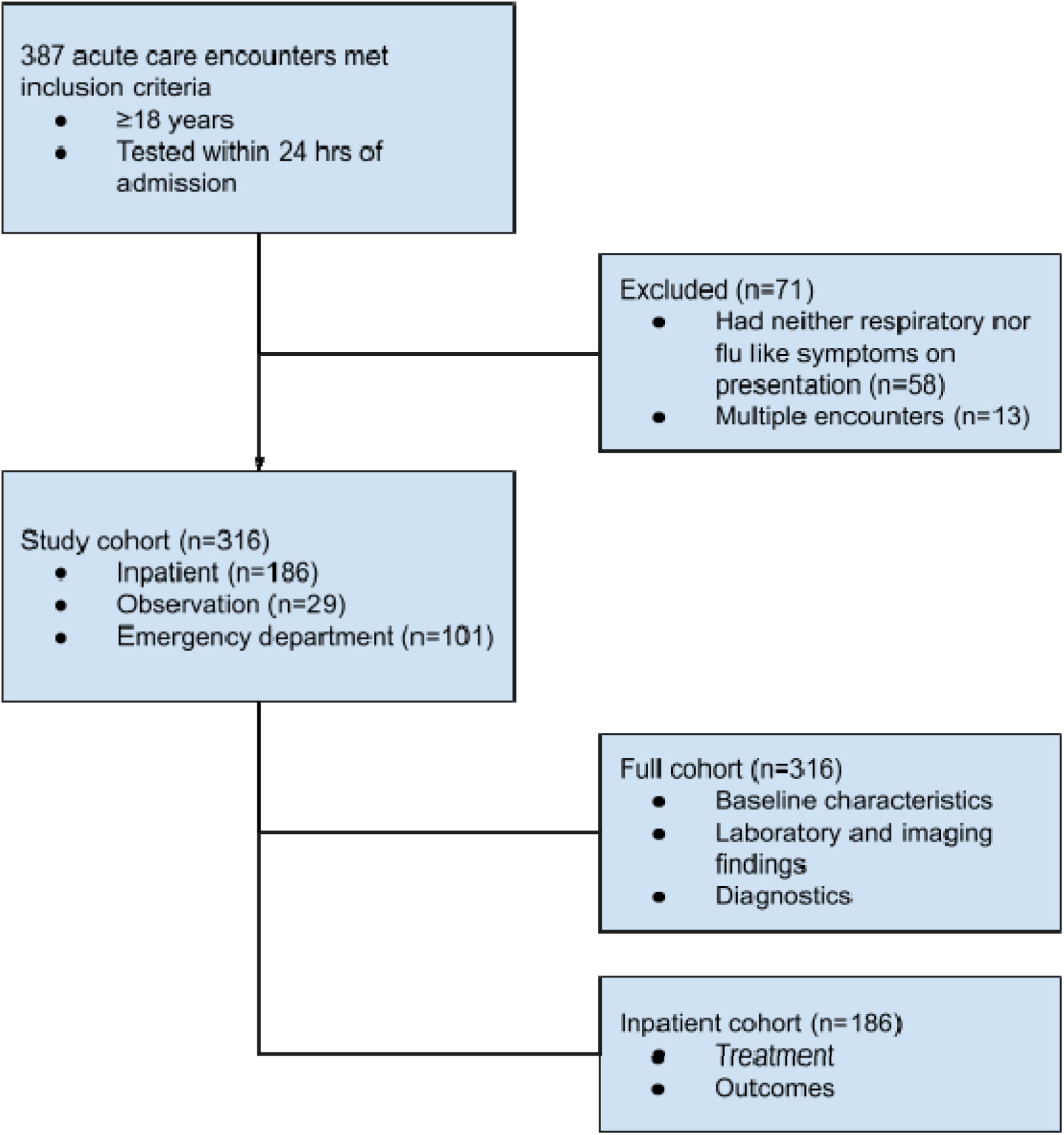
Cohort flow diagram.

**Appendix Table 1:** Results of chest CT performed within 24 hours of admission

| **Chest CT findings** | COVID-19 positive (n=33) | COVID-19 negative (n=283) | P value |
| --- | --- | --- | --- |
| CT within first 24 hours | 2/33 (6%) | 60/283 (21%) | 0.038 |
| Focal consolidation<br>Unilateral<br>Bilateral<br>Not Present | 0/2 (0%)<br>2/2 (100%)<br>0/2 (0%) | 15/60 (25%)<br>9/60 (15%)<br>36/60 (60%) | 0.029 |
| Ground-glass opacities<br>Unilateral<br>Bilateral<br>Not Present | 0/2 (0%)<br>2/2 (100%)<br>0/2 (0%) | 7/60 (12%)<br>19/60 (32%)<br>34/60 (58%) | 0.200 |
| Septal thickening<br>Unilateral<br>Bilateral<br>Not Present | 0/2 (0%)<br>0/2 (0%)<br>2/2 (100%) | 3/60 (5%)<br>10/60 (17%)<br>47/60 (78%) | 1.000 |
| Pleural effusion<br>Unilateral<br>Bilateral<br>Not Present | 0/2 (0%)<br>0/2 (0%)<br>2/2 (100%) | 6/60 (10%)<br>11/60 (18%)<br>43/60 (72%) | 1.000 |
| Lymphadenopathy | 1/2 (50%) | 15/60 (25%) | 0.453 |

**Appendix Table 2:** Treatment of Emergency department and observation patients with COVID19 infection

|  | COVID positive (n=7) | COVID negative (n=123) | P Value |
| --- | --- | --- | --- |
| **Treatment** |  |  |  |
| Doxycycline | 2/7 (29%) | 13/123 (11%) | 0.186 |
| Fluoroquinolones | 0/7 (0%) | 3/123 (2%) | 1.00 |
| Azithromycin | 2/7 (29%) | 4/123 (3%) | 0.033 |
| Cephalosporin | 1/7 (14%) | 4/123 (3%) | 0.245 |
| TMP-SMX | 0/7 (0%) | 2/123 (2%) | 1.00 |
| Oseltamivir | 0/7 (0%) | 4/123 (3%) | 1.00 |
| No antimicrobials given on dc | 3/7 (43%) | 100/123 (80%) | 0.041 |
| Respiratory support |  |  |  |
| Supplemental oxygen<br>High Flow | 0/7 (0%)<br>0/7 (0%) | 3/123 (3%)<br>0/123(0% ) | 1.00<br>-- |
| Crystalloid bolus volume within first 24<br>hours (mean, SD) | 1000 (0)<br>n=3 | 1351.4 (716)<br>n=37 | 0.406 |

**Appendix Figure 2:**
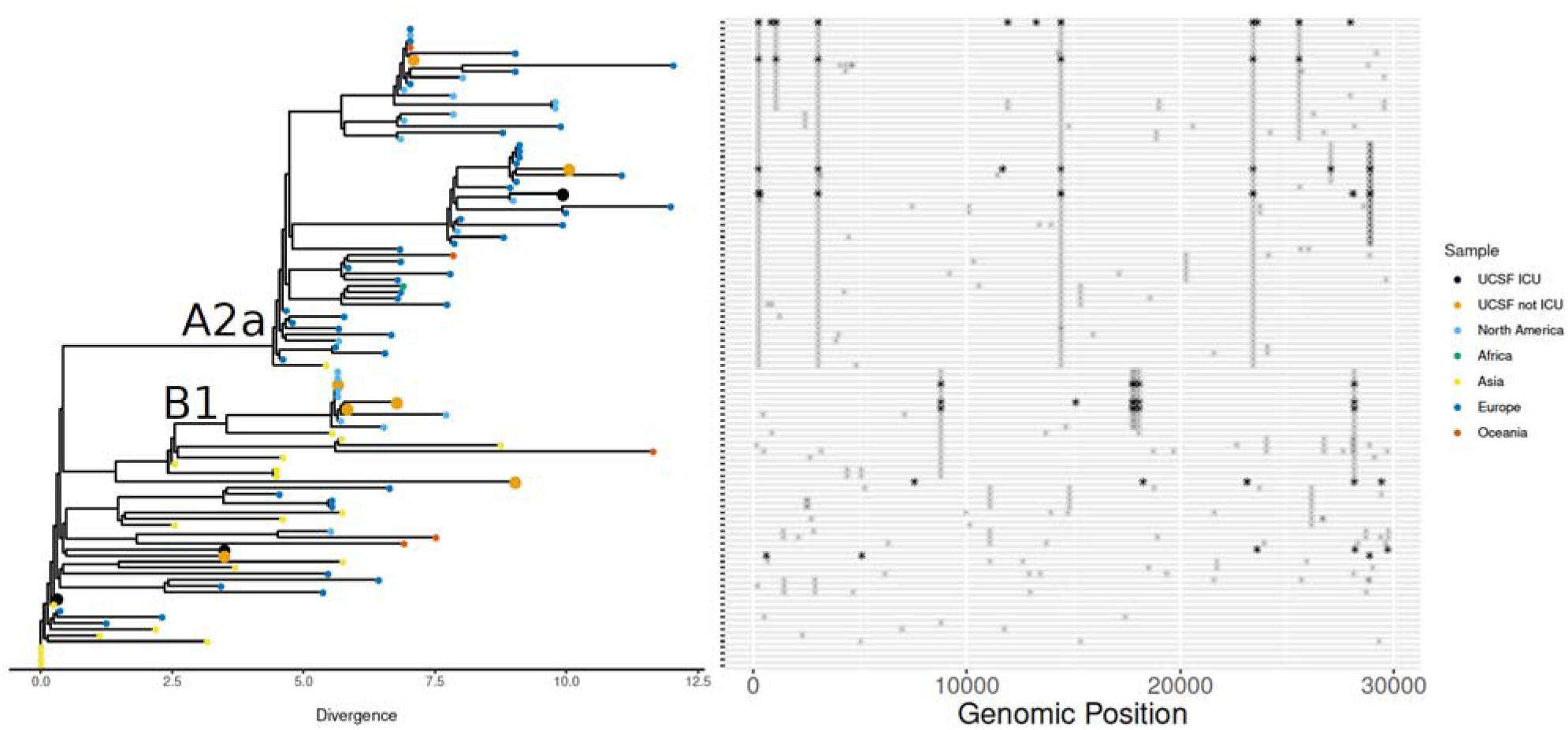
Genomic epidemiology of SARS-CoV-2 in study population. Phylogenetic analysis of 10 SARS-CoV-2 genomes from patients in the cohort indicated strains originating from a diversity of geographic locations. Single nucleotide polymorphisms are plotted in the panel adjacent to the phylogenetic tree. Most samples fell into the Nextstrain.org clades A2a (widely prevalent in New York) and B1 (detected in Washington State in February 2020). The SARS-CoV-2 from patients who required ICU care were not associated with any single clade.

**Appendix Table 3:** Complete microbiological test results for each patient. Legend: Respiratory culture: sputum, endotracheal aspirate or bronchoalveolar lavage; negative: not detected; n/a = not applicable because RNA from patient sample unavailable for testing; invalid = sample unable to be analyzed by mNGS due to insufficient (<25pg) RNA.

